# SARS-CoV-2 variant Delta rapidly displaced variant Alpha in the United States and led to higher viral loads

**DOI:** 10.1101/2021.06.20.21259195

**Authors:** Alexandre Bolze, Elizabeth T. Cirulli, Shishi Luo, Simon White, Dana Wyman, Andrew Dei Rossi, Henrique Machado, Tyler Cassens, Sharoni Jacobs, Kelly M. Schiabor Barrett, Kevin Tsan, Jason Nguyen, Jimmy M. Ramirez, Efren Sandoval, Xueqing Wang, David Wong, David Becker, Marc Laurent, James T. Lu, Magnus Isaksson, Nicole L. Washington, William Lee

## Abstract

This study reports on the displacement of Alpha (B.1.1.7) by Delta (B.1.617.2 and its substrains AY.1, AY.2, and AY.3) in the United States. By analyzing RT-qPCR testing results and viral sequencing results of samples collected across the United States, we show that the percentage of SARS-CoV-2 positive cases caused by Alpha dropped from 67% in May 2021 to less than 3.0% in just 10 weeks. We also show that the Delta variant has outcompeted the Iota (B.1.526) variant of interest and Gamma (P.1) variant of concern. An analysis of the mean quantification cycles (Cq) values in positive tests over time also reveal that Delta infections lead to a higher viral load on average compared to Alpha infections, but this increase is only 2 to 3x on average for our study design. Our results are consistent with the hypothesis that the Delta variant is more transmissible than the Alpha variant, and that this could be due to the Delta variant’s ability to establish a higher viral load earlier in the infection compared to the Alpha variant.

## Introduction

The SARS-CoV-2 Delta variant, which includes variants B.1.617.2, AY.1, AY.2 and AY.3 has been classified as a variant of concern (VOC) by Public Health England (PHE), the World Health Organization (WHO), and the U.S. Centers for Disease Control (CDC) ^1^. The Delta variant was the predominant variant in India during the peak in cases in April and May 2021^2^. When it spread to England, the Delta variant led to a new wave of cases and outcompeted the then-dominant Alpha variant, also named B.1.1.7 ^3,4^. By July 2021, it had been identified in 112 countries ^2^. The Delta variant has been shown to be more transmissible than the SARS-CoV-2 Alpha variant in England ^3^. This higher transmissibility is not thought to be due to a decrease of vaccine effectiveness against Delta; a study by Public Health England showed that vaccine efficacy for AstraZeneca and Pfizer vaccines remained very good (>90%) against hospitalizations after two doses ^5^. Instead, a study found that viral load increases rapidly early in a Delta infection compared to the original SARS-CoV-2 strain, suggesting a shorter incubation period, faster viral replication, and increased infectiousness earlier in infection, all leading to a higher basic reproductive ratio^6^.

The context in the United States is different from England or India. In addition to differences in public health policies and vaccination rates, the variants that were co-circulating when Delta was introduced also differed between England and the United States^2^. In England, Alpha represented more than 90% of the SARS-CoV-2 sequences when Delta was first identified in the country (around March 2021), and there were very few sequences of Gamma, also named P.1, another variant of concern. In the United States, Alpha had plateaued just above 70%, and there was a greater diversity of variants when Delta started to emerge (also around March 2021), including an increasing amount of Gamma^7^, and about 10% of Iota, another variant of interest^8^.

The objectives of this study are therefore (i) to analyze the impact of the introductions of the Delta variant on the prevalence of Alpha in the United States, and (ii) to test the hypothesis that the higher transmissibility of Delta is due to a higher viral load in the nose. To this end, we looked at the RT-qPCR testing results and sequencing results of samples collected by the Helix laboratory across the United States in 2021. Importantly, the collection method and collection sites have been consistent since February 2021, and the samples analyzed should not be biased for localized outbreaks. We therefore make the assumption that there was no significant sampling bias between the testing and sequencing done by our lab from February to April 2021 when Alpha was rapidly increasing in the United States, and the months of May, June and July 2021.

## Methods

### Ethical statement

The Helix data analyzed and presented here were obtained through IRB protocol WIRB#20203438, which grants a waiver of consent for a limited dataset for the purposes of public health under section 164.512(b) of the Privacy Rule (45 CFR § 164.512(b)).

### Helix COVID-19 test data and sample selection

All viral samples in this investigation were collected by Helix through its COVID-19 diagnostic testing laboratory. The Helix COVID-19 Test (EUA 201636) was run on specimens collected across the US, and results were obtained as part of our standard test processing workflow using specimens from anterior nares swabs. The Helix COVID-19 Test is based on the Thermo Fisher TaqPath COVID-19 Combo Kit, which targets three SARS-CoV-2 viral regions (N gene, S gene, and ORF1ab). Test results from positive cases, together with a limited amount of metadata (including sample collection date, state, and RT-qPCR Cq values for all gene targets), were used to build the research database used here. Ongoing summary level data are viewable at https://www.helix.com/covid19db.

For this study, we analyzed 377,514 positive samples collected from January 1 2021 to July 27 2021. We collected samples from all 50 states of the United States, though not necessarily in proportion to each state’s population. **Table 1** shows the ten states for which we have the most samples in this study.

**Table 1.** Counts of positive samples collected in 2021 and analysed in this study by state.

| state | counts positive in 2021 |
| --- | --- |
| FL | 113,942 |
| CA | 59,968 |
| PA | 39,633 |
| GA | 31,642 |
| TX | 25,973 |
| NC | 20,795 |
| MI | 20,440 |
| IN | 19,781 |
| MA | 15,128 |
| AZ | 10,301 |
| other states | 19,911 |

### SARS-CoV-2 sequencing and consensus sequence generation

Sequencing was performed by Illumina^9^, and by Helix since June 2021, as part of the SARS-CoV-2 genomic surveillance program led by the Centers for Disease Control and Prevention (CDC). In the Helix workflow, RNA was extracted from 400 μl of patient anterior nares sample using the MagMAX Viral/Pathogen kit (ThermoScientific). All samples were subjected to total RNA library preparation using the Rapid RNA Library Kit Instructions (Swift Biosciences). SARS-COV-2 genome capture was accomplished using hybridization kit xGen COVID-19 Capture Panel (Integrated DNA Technologies). Samples were sequenced using the NovaSeq 6000 Sequencing system S1 flow cell, which included the NovaSeq 6000 Sequencing System S1 Reagent Kit v1.5 (300 cycles). Bioinformatic processing of this sequencing output was as follows. The flow cell output was demultiplexed with bcl2fastq (Illumina) into per-sample FASTQ sequences that were then run through the Helix klados-fastagenerator pipeline v1.6.0 to produce a sequence FASTA file. First, reads were aligned to a reference comprising the SARS-CoV-2 genome (NCBI accession NC_045512.2) and the human transcriptome (GENCODE v37) using BWA-MEM. Following duplicate-marking, SARS-CoV-2 variants were called using the Haplotyper algorithm (Sentieon, Inc). The per-base coverage from the alignment file (BAM) and per-variant allele depths from the variant call format (VCF) file were then used to build a consensus sequence according to the following criteria: if there are at least 5 unique reads covering a base, and at least 80% of the reads support a particular allele, that allele is reported. Otherwise, that base is considered uncertain, and an N is reported.

### Viral lineage designation

Viral sequences were assigned a Pango lineage ^10^ using pangoLEARN (https://github.com/cov-lineages/pangoLEARN). For this analysis, pangoLEARN version 2021-07-09 with Pangolin software version 3.1.7 was used. We sequenced and were able to attribute a lineage to 56,308 sequences from samples collected in 2021.

### Cq analysis

We restricted our analysis of Cq values to samples that had both N gene Cq and ORF1ab Cq < 35 (n=347,187 samples). This was to minimize a potential bias driven by samples with very low viral load that are more likely to have one of the three targets drop, leading to an artificially higher mean Cq for SGTF.

Comparison of Cqs between different weeks were done using an unpaired t-test with PRISM version 8.

## Results

### The rapid decrease of Alpha in the US

One of the defining mutations of the Alpha variant of concern is the deletion of amino acids 69 and 70 in the spike protein. This deletion interferes with the PCR test target on the S gene in many COVID-19 tests ^11^, including the Helix COVID-19 Test, and causes S-gene target failure (SGTF). In January 2021, SGTF positives were found to be caused by Alpha variants, as well as a few other variants such as B.1.375. Moreover, the S-gene target may fail if viral load is low and Cq is high, usually above 35. To assess whether SGTF could be used to study the increase or decrease of the Alpha variant of concern in the United States, we looked at all 5,344 sequences from SGTF samples in May, June and July 2021. Of those, 99.3% (5,306 of 5,344) were Alpha (**Figure 1A**). The next lineage leading to SGTF in that time period in the United States was Eta, also named B.1.525, (12 of 5,344). Of note, there were two SGTF samples sequenced that were Gamma. The other Gamma variants sequenced, as well as all other variants of concern that are not Alpha, did not lead to SGTF. And 96.4% (5,306 of 5,505) of the sequences assigned an Alpha lineage in May, June and July 2021 were SGTF. SGTF is therefore a reliable proxy to track the epidemiological dynamics of Alpha in the United States.

**Figure 1:**
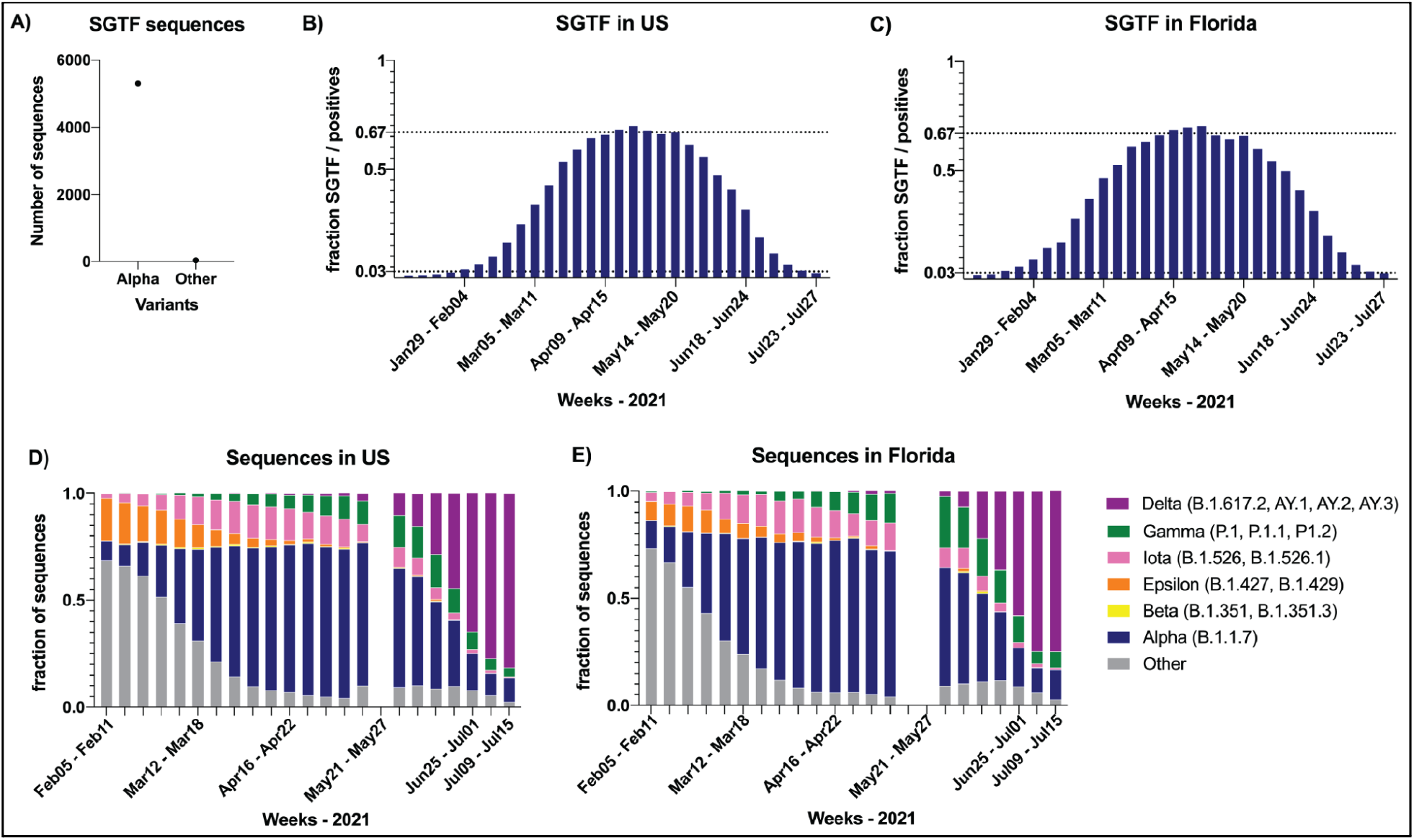
Alpha variant was replaced by Delta variant in the United States. A) Counts of S-Gene Target Failure (SGTF) sequenced in May, June and July 2021 that were Alpha or Other variants. B) Fraction of SGTF to total positives per week in the US. C) Fraction of SGTF to total positives per week in Florida. D-E) Fraction of sequences by lineage per week in the US (panel D) and in Florida (panel E). Purple: Delta (B.1.617.2, AY.1, AY.2, AY.3). Green: Gamma (P.1, P.1.1, P.1.2)). Pink: Iota (B.1.526, B.1.526.1, B.1.526.2). Orange: Epsilon (B.1.427, B.1.429). Yellow: Beta (B.1.351, B.1.351.3). Blue: Alpha (B.1.1.7). Other lineages are grouped together in gray.

We therefore analyzed 274,591 positive samples for SARS-CoV-2 with a Cq for the N gene <29 and tested at the Helix laboratory between January 1 and July 27 2021. Both SGTF and sequencing data indicate that the Alpha variant, after becoming the dominant SARS-CoV-2 lineage in the United States ^9,12^ plateaued at around 70% prevalence at the end of April 2021 (**Figure 1B**). By looking at May, June and July test results in the US, we saw a clear and rapid decrease of the fraction of SGTF among positive results, decreasing from 67% in week 20 (May 14 - 20) to 2.3% in week 30 (July 23 - 27) of 2021 (**Figure 1B**). To ensure this result was not driven by a change in the states or regions with a high number of cases, or other artefacts, we examined the trend in Florida alone and observed the same rapid decrease in the fraction of SGTF: from 66% of positives in week 20 to only 2.8% in week 30 (**Figure 1C**). Overall, these results show that the variant of concern Alpha was rapidly displaced in the United States between May and July 2021.

### The Delta variant was responsible for the decrease of the Alpha variant

We analyzed the Pango lineage associated with each sequence to investigate which variants might be displacing Alpha in the United States. While the fraction of Gamma sequences grew in April and May 2021, it plateaued at 15% of sequences by week 22 of 2021 (May 28 to June 03) in the US, even as Alpha continued to decline in prevalence (**Figure 1D**). We observed the same in Florida where Gamma grew to represent up to 24% of the sequences in Florida in week 22 of 2021 (May 28 to June 03) before decreasing, too (**Figure 1E**). By week 28 of 2021 (July 09 to July 15), Gamma represented only 4.3% (10 of 234) of sequences in the US. The only lineage that steadily grew from April to July 2021 was Delta (**Figure 1D** - **1E**). By week 28 of 2021, the Delta lineages represented 81.6% (191 of 234) of the sequences in our dataset (**Figure 1D**). The same was true when restricting the analysis to Florida, where Delta represented 75% of the sequences by week 28. Given the continued decrease of SGTF at weeks 29 and 30, we expect that Delta will account for about 90% of the sequences by week 30 in the United States. Overall, these results show that the Delta variant displaced the Alpha variant in the United States in the summer of 2021, and, while doing so, also outcompeted other variants of concern (Gamma) and variants of interest (Iota).

### Higher viral load in cases due to Delta compared to Alpha and other S-positive variants

We next tried to understand what could be driving this advantage of Delta compared to Alpha and other variants. One hypothesis is that Delta virions replicate more rapidly and to higher viral loads in the nose, so that individuals exposed to the Delta variant become infectious earlier, compared to individuals exposed to other variants. In a study of quarantined subjects, researchers observed that subjects infected with the Delta variant had detectable virus (by RT-qPCR) two days earlier and at a 1000-fold higher titer (∼10 Ct, cycles threshold) than subjects infected with the initial SARS-CoV-2 virus from early 2020^6^. We therefore tested the hypothesis that we would observe higher viral loads, corresponding to lower average Cq values, in infections by Delta compared to infections by Alpha or other variants. One difference in our study design, compared to the earlier study, was that we did not know the time between when a patient had a detectable virus and when the patient was tested. Given that individuals are more likely to test if they’re symptomatic, and given that viral titers are typically declining by symptom onset^13^, it is likely our tests measure viral loads several days after a patient has had detectable virus. Nonetheless, as described below, our large sample size and the fact that all samples were processed in the same laboratory allowed us to look at the trends of Cq numbers over time based on the abundance of different variants.

We split our cohort into SGTF and S-positive samples, calculating the mean Cq levels over time for each group separately. By mid-February, >90% of SGTF samples were Alpha^9^, and we leverage this fact to make the assumption below that Cq values of SGTF samples are representative of Cq values in infections caused by the Alpha variant. We observed that while the mean Cq level for SGTF samples was lower (0.5 to 1 Cq lower) compared to S-positive samples every week from week 3 to week 20, this trend started to invert after week 20, around the time where Delta accounted for more than 10% of the S-positive samples (**Figure 2A**). From week 20 to week 30, the mean Cq level for S-positive samples decreased compared to mean Cq level for SGTF samples (**Figure 2A**) suggesting that S-positive samples were exhibiting higher viral loads. When focusing on the evolution of the mean Cq level of S-positive samples from week 17 to 30, we observed a linear decrease correlating with the increase in the fraction of Delta samples calculated in our previous analysis (**Figure 2B**).

**Figure 2:**
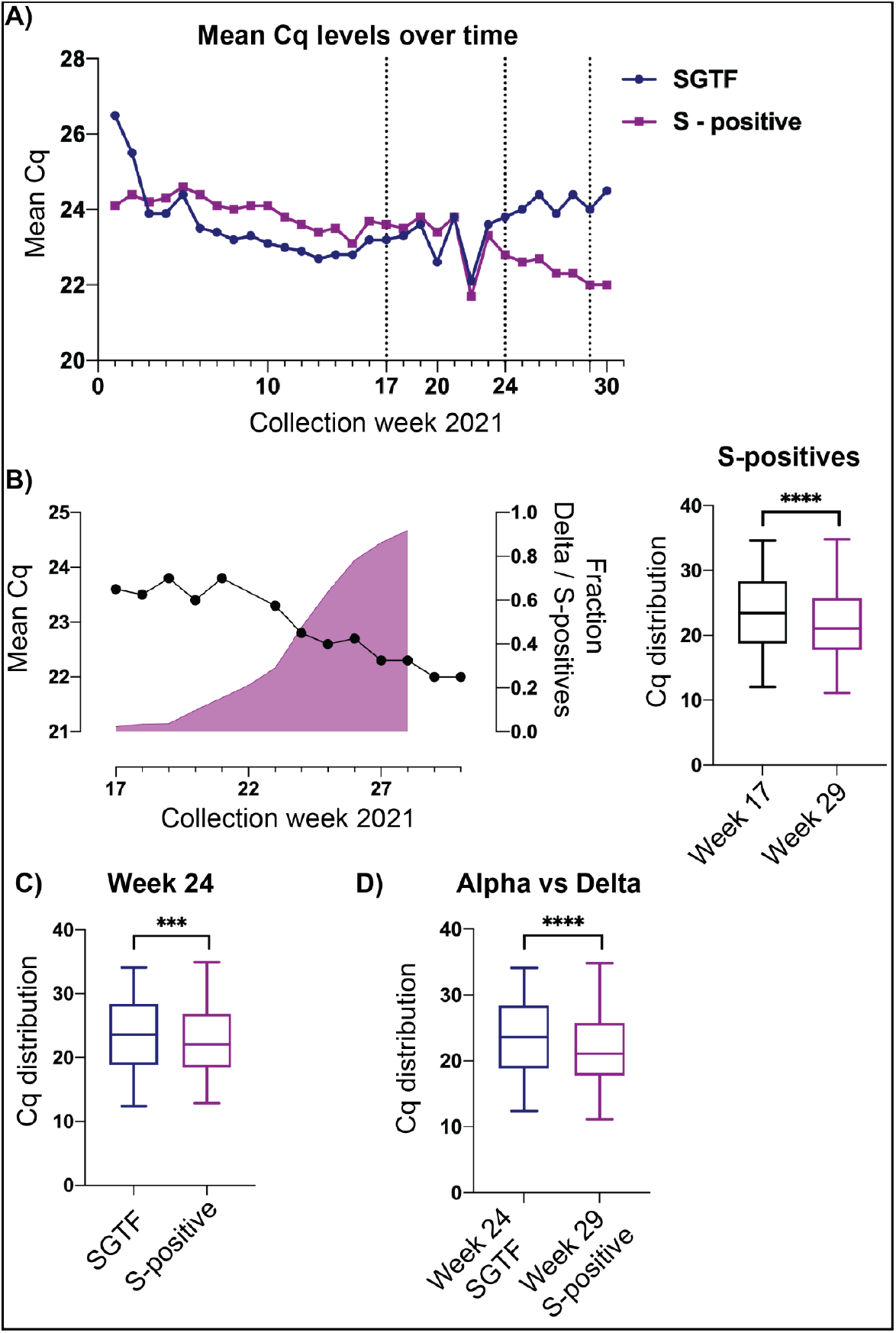
Higher viral loads in Delta infections compared to Alpha and other variants. A) Trend of mean Cq levels over time in 2021. Each dot corresponds to a week. Blue line represents positives with SGTF, a great proxy for Delta variant. Purple line represents S-positives. The three vertical dotted lines indicate weeks 17, 24 and 29, which are later analyzed in the panels below. B) Left panel zooms in on the trend of the mean Cq level for S-positive samples between weeks 17 to 30. The purple graph on the left panel indicates the fraction of S-positives that are the Delta variant. The right panel are box plots comparing distributions of Cq levels between week 17 and 29 (in purple because mostly Delta variant). C) Distribution of Cq levels at week 24. Blue box is SGTF. Purple box is S-positive. D) Distribution of Cq levels for SGTF at week 24 (blue box) and S-positive at week 29 (purple box). Unpaired t-tests were performed. ****: p<0.0001, ***: p<0.001.

To compare the mean Cq level for Delta infections with infections from other S-positive variants, we compared the Cq levels from all S-positives at week 17 (almost no Delta) with the Cq levels at week 29 (>90% Delta) (**Figure 2B**). The results showed a difference of 1.62 Cq (95% CI: 1.41 - 1.84, p<0.0001 unpaired t-test) (**Figure 2B**). To compare the viral loads of Alpha with Delta, we decided to focus on recent weeks when SGTF represented about half of the samples while the Delta variant was increasing. The objective was to mitigate possible biases due to the surge of cases and overall public messaging on testing. We first performed the comparison of both groups at week 24, where about half of all positives were SGTF (>99% of those are Alpha), and about half of the S-positives are Delta. The results showed a difference of 0.93 Cq (95% CI: 0.45 - 1.42, p=0.0002 unpaired t-test) (**Figure 2C**). We then compared the Cqs from SGTF of week 24 (>99% Alpha) with the Cqs from S-positives of week 29 (>90% Delta), which is closer to a direct comparison between Alpha and Delta. The results showed a difference of 1.76 Cq (95% CI: 1.39 - 2.14, p<0.0001 unpaired t-test) (**Figure 2D**). Overall, our analysis of Cq results from RT-qPCR tests showed that on average, in a large sample size, the viral loads from Delta infections was ∼2.5 to 3 times higher (1.6 to 1.8 Cq) compared to Alpha and the other main variants present in the United States in May and June 2021.

## Discussion

Here, we use RT-qPCR results from 377,514 positive samples and 56,308 viral sequences from Helix COVID-19 tests collected since January 2021 to show the trajectories of different variants of concern in the United States. The total percentage of positive COVID-19 tests attributed to Alpha in the United States fell from a peak of 70% in April down to less than 3% in the most recent week (3rd week of July 2021). We showed that most of the displacement of Alpha can be attributed to Delta. Our results are consistent with those from Public Health England^3^. In addition, we showed that Delta also displaced Gamma and Iota in the United States, viral competition dynamics that could not be studied in England.

Our analysis of the Cq (quantification cycle) is consistent with the hypothesis that the Delta variant can replicate faster and lead to higher viral loads earlier in infection compared to other variants. The difference we observed of 2.5 to 3x higher viral load was small compared to the ∼1000x difference observed in the study of quarantined individuals^6^. However the study designs were vastly different. First, Li et al compared Delta infections to infections with the earliest form of the virus (19A/B) present in China in early 2020. It is very likely that the S:D614G mutation and other recent mutations acquired since have also led to a faster replication of the virus. The difference in viral load may be less than 1,000x if the comparison was between Delta infections and Alpha infections. Secondly, the samples studied here came from a community setting, with the large majority from a national retail pharmacy. For the majority of these samples, it is unlikely that they were collected right after the time of infection. An additional limitation to our analysis is that samples are collected from multiple locations, which could add further variables that affect the viral load in each sample.

Overall our analysis showed that the SARS-CoV-2 Delta variant displaced the Alpha variant in the United States between May and July 2021. One biological explanation to this could be that Delta replicates faster and Delta infections lead to higher viral loads earlier after infection. Additional studies looking at viral loads in a large number of samples like ours, or in a more controlled study design are still needed to understand the time period when the majority of individuals infected by Delta are infectious. Other studies focused on Delta infections in fully vaccinated individuals will be of interest, too, to understand the kinetics of Delta replication in these individuals. Many additional questions about the rise of the Delta variant remain unexplained. One of them is to understand why the Delta variant spread and took over England 3 to 4 weeks before the United States despite a similar introduction based on the current first sequences made public^2^.

## Data Availability

Data used in this manuscript are available from the Data tables in supplementary material. It can also be accessed on Helix Github COVID page: https://github.com/myhelix/helix-covid19db. Sequences are also available on GISAID.

## Acknowledgements

We thank the employees of Helix, employees of Illumina, members of the CDC SPHERES consortium and California CovidNET, and members of the Andersen Lab at Scripps Research for discussion and help with logistics. We thank the healthcare workers, frontline workers, and patients who made the collection of this SARS-CoV-2 dataset possible. This work has been funded in part by CDC BAA contract 75D30121P10258 (Illumina, Helix). We also want to give a special thanks to Public Health England for their inspiring work and technical reports, as well as the team behind outbreak.info, a resource that we used throughout this study.

